# SARS-CoV-2 has been circulating in northern Italy since December 2019: evidence from environmental monitoring

**DOI:** 10.1101/2020.06.25.20140061

**Authors:** Giuseppina La Rosa, Pamela Mancini, Giusy Bonanno Ferraro, Carolina Veneri, Marcello Iaconelli, Lucia Bonadonna, Luca Lucentini, Elisabetta Suffredini

**Affiliations:** Department of Environment and Health, Istituto Superiore di Sanità, Rome, Italy; Department of Food Safety, Nutrition and Veterinary Public Health, Istituto Superiore di Sanità, Rome, Italy

**Keywords:** SARS-CoV-2, Coronavirus, COVID-19, sewage, wastewater, surveillance

## Abstract

Severe Acute Respiratory Syndrome Coronavirus 2 (SARS-CoV-2) is responsible for the coronavirus disease COVID-19, a public health emergency worldwide, and Italy is among the world’s first and most severely affected countries. The first autochthonous Italian case of COVID-19 was documented on February 21. We investigated the possibility that SARS-CoV-2 emerged in Italy earlier than that date, by analysing 40 composite influent wastewater samples collected - in the framework of other wastewater-based epidemiology projects - between October 2019 and February 2020 from five wastewater treatment plants (WTPs) in three cities and regions in northern Italy (Milan/Lombardy, Turin/Piedmont and Bologna/Emilia Romagna). Twenty-four additional samples collected in the same WTPs between September 2018 and June 2019 were included as blank samples. Viral concentration was performed according to the standard World Health Organization procedure for poliovirus sewage surveillance. Molecular analysis was undertaken with both nested RT-PCR and real-rime RT-PCR assays. A total of 15 positive samples were confirmed by both methods. Of these, 8 were collected before the first autochthonous Italian case. The earliest dates back to 18 December 2019 in Milan and Turin and 29 January 2020 in Bologna. Samples collected in January and February in the three cities were also positive.

Our results demonstrate that SARS-CoV-2 was already circulating in northern Italy at the end of 2019. Moreover, it was circulating in different geographic regions simultaneously, which changes our previous understanding of the geographical circulation of the virus in Italy. Our study highlights once again the importance of environmental surveillance as an early warning system, to monitor the levels of virus circulating in the population and identify outbreaks even before cases are notified to the healthcare system.

## INTRODUCTION

Coronaviruses (CoVs) belong to the Coronaviridae family and are enveloped, single-stranded RNA viruses, grouped into four main groups: alpha, beta, gamma and delta CoVs. Most human coronaviruses cause mild respiratory infections (CoV 229E, NL63, OC43, and HKU1). Some CoVs, however, are associated with severe symptoms and outbreaks. These are MERS-CoV (the beta coronavirus that causes Middle East Respiratory Syndrome, or MERS), SARS-CoV (the beta coronavirus that causes severe acute respiratory syndrome, or SARS), and the recently discovered SARS-CoV-2 (the novel coronavirus that causes coronavirus disease 2019, or COVID-19).

SARS-CoV-2 was discovered in December 2019 in China, and has then spread widely in many countries, to the point that, on 11 March 2020, the World Health Organization (WHO) declared COVID-19 a pandemic. Italy has been among the first, and most severely affected countries in the world - as of 15 June 2020, 237,695 COVID-19 cases were diagnosed, with 33,168 deaths (https://www.epicentro.iss.it/coronavirus/bollettino/Infografica_15giugno%20ITA.pdf). However, it is likely that, in Italy as well as in all other affected countries in the world, the true number of cases has been substantially greater than reported, as mild or asymptomatic infections have often been overlooked.

The first SARS-CoV-2 cases reported in Italy were two Chinese tourists who fell ill in January after flying in from Wuhan, where the epidemic began. These patients were immediately put into isolation, and are not believed to have infected anyone else. The first autochthonous patient was diagnosed one month later in Lombardy, on February 21. He was a 38-year-old man, from the town of Codogno, 60 km southeast of Milan. Initially, it was believed that “patient zero” might have been a colleague of his who had recently returned from a business trip to China. This colleague tested negative, however, so the first introduction of the virus into Italy remains unclear.

Identifying the first introduction of the virus is of great epidemiological interest. In Italy, and elsewhere, there have been speculations to the effect that COVID-19 had been silently circulating before the first case was identified. Indeed, other countries have been trying to ascertain whether earlier infections had occurred. In France, where the COVID-19 epidemic was believed to have started in late January 2020, a retrospective analysis of a stored respiratory sample from a patient hospitalised in December 2019, demonstrated that the patient was positive for SARS-CoV-2, suggesting that, in France, the epidemic started much earlier than previously thought (Deslandes *et al*., 2020).

It is known that gastrointestinal symptoms are seen in patients with COVID-19 (between 16% to 33% in most studies), and that approximately 50% of patients with COVID-19 have detectable virus in their stool (Ouali et al., 2020). These patients have been shown to shed the virus in their stools even if asymptomatic or pre-symptomatic (Jiang *et al*., 2020;Park *et al*., 2020;Tang *et al*., 2020). Sewage samples can thus be used to monitor the levels of virus circulating in the population, an approach called wastewater-based epidemiology (WBE). Several studies performed in the Netherlands (Medema *et al*., 2020), the United States (Wu *et al*., 2020; Nemudryi et al., 2020), France (Wurtzer et al., 2020), Australia (Ahmed *et al*., 2020a), Spain (Randazzo *et al*., 2020;Chavarria-Mirò *et al*., 2020), Japan (Hata *et al*., 2020), Turkey (Kocamemi et al., 2020), and Israel (Bar-Or et al., 2020) have demonstrated that sewage surveillance can help understand the circulation of SARS-CoV-2 in human populations. In Italy, our group has previously found SARS-CoV-2 in sewage samples collected between the end of February (after the first autochthonous case) and April 2020 (La Rosa *et al*., 2020). Another Italian study confirmed the occurrence of the virus in sewage samples collected in April (Rimoldi *et al*., 2020). Thus far, all of the cited studies performed worldwide, have analysed wastewater samples collected during the pandemic, with the exception of the Spanish study of Chavarria-Mirò and co-worker, who also analysed frozen archival samples from 2018 (January-March) and 2019 (January, March, September-December) Chavarria-Mirò *et al*., 2020). Similarly, in this study we retrospectively searched for genomic traces of SARS-CoV-2 in a collection of sewage samples gathered from WTPs in northern Italy between October 2019 and February 2020, in the framework of different WBE projects on enteric viruses. The samples were analysed to ascertain whether SARS-CoV-2 was circulating in the weeks and months before the virus was believed to have arrived in Italy.

## MATERIALS AND METHODS

### Sampling and sample preparation

Forty sewage samples were analysed for the study. Samples were collected between 9 October 2019 and 28 February 2020 from five WTPs, located in Milan (20 samples from two distinct plants, referred to as A and B), Turin (16 samples from plants C and D), and Bologna (4 samples from plant E). The location and number of inhabitants (expressed as population equivalents) served by these WTPs are summarised in Figure 1. Other 24 wastewater samples, collected from the same WTPs in Milan, Turin and Bologna between 12 September 2018 and 19 June 2019 (i.e. before the emergence of SARS-CoV-2 as a human pathogen), were analysed as ‘blank samples’.

**Figure 1:**
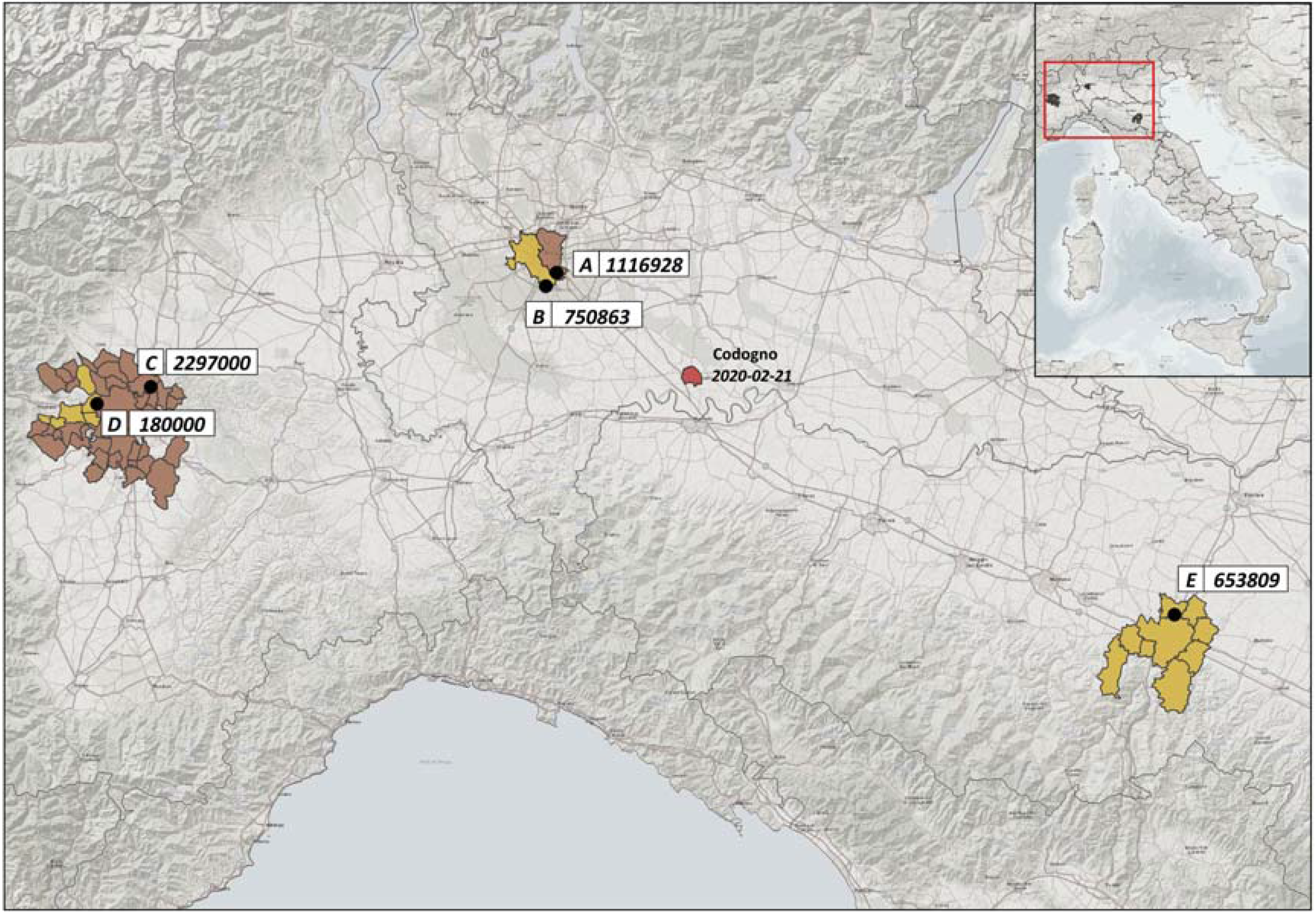
Location and number of inhabitants served by the WTPs included in the study. Numbers in correspondence of the WTP code represent the inhabitants served by each plant

Composite samples, representing 24-hour periods, were collected raw, before treatments, stored at - 20 °C, and dispatched frozen to Istituto Superiore di Sanità (the Italian National Institute of Health) for analysis. Precautions taken during sample treatment were reported elsewhere (La Rosa et al., 2020). Before sample concentration, a 30 min viral inactivation treatment at 56 °C was undertaken in order to increase the safety of the analytical protocol for both laboratory personnel and the environment. Sample concentration was performed using the two-phase (PEG-dextran) separation method recommended by the WHO Guidelines for environmental surveillance of poliovirus circulation (WHO, 2003), with modifications. Briefly, 250 mL of wastewater sample was centrifuged (30 min at 1200 × *g*) to separate the pellet. The pellet was kept at 4 °C to be later combined with the concentrated supernatant. The clarified wastewater was neutralized (pH 7.0-7.5), mixed with dextran and polyethylene glycol (19.8 ml of 22% dextran, 143.5 ml 29% PEG 6000, and 17.5 ml 5N NaCl), and after a constant agitation for 30 minutes using a horizontal shaker, the mixture was left to stand overnight at 4 °C in a separation funnel. Viruses, accumulated in the smaller bottom layer and/or at the boundary between the layers (interphase), were then collected drop-wise, and this concentrate was re-joined to the pellet retained after the initial centrifugation. In a previous study by our group on SARS-CoV-2 detection in sewage, the original WHO protocol was modified by omitting the chloroform treatment after collecting the concentrate, to avoid loss of SARS-CoV-2 particles, since lipid-containing viruses are chloroform sensitive (La Rosa et al., 2020). However, RNA obtained from those samples were found to be moderately inhibited (median inhibition 29.1%, range 8.7% - 51.4%). Therefore, after performing comparative extraction experiments with and without chloroform, using field samples and samples spiked with the human Alphacoronavirus HCoV 229E (data not shown), the chloroform purification step was reintroduced to improve the purification of samples before RNA extraction, and obtain a higher detection sensitivity. The concentrated sample was then extracted with 20% (v/v) of chloroform by shaking vigorously for 1 min and centrifugation (1400 × *g* for 10 min). The total recovered volume (ranging from 7 to 10 ml) was then recorded, and half of the concentrate was subjected to genome extraction, the remaining being stored at −80 °C.

The recovery efficiency of the concentration and extraction procedure was assessed through separate spiking experiments performed in quadruplicate using the Alphacoronavirus HCoV 229E (ATCC VR-740). This was not done on field samples in order to avoid interferences with future virome analyses.

Genome extraction was performed using the NucliSENS miniMAG semi-automated extraction system with magnetic silica (bioMerieux, Marcy l’Etoile, France), with the following modifications to the manufacturer’s protocol to adapt to large volumes: the quantity of lysis buffer added was the equivalent of twice the volume of the sample, the lysis phase was prolonged to 20 minutes, and 100 μl magnetic silica beads were used per sample. The subsequent washing phases were performed as per manufacturer’s instructions. Before molecular tests, extracted RNAs were purified from residual PCR inhibitors using the OneStep PCR Inhibitor Removal Kit (Zymo Research, CA, USA).

### Nested RT-PCR

RNAs were tested for the presence of SARS-CoV-2 by the nested RT-PCR assays in the ORF1ab region (Table 1) used to detect the first positive sewage samples in Italy (La Rosa et al., 2020).

**Table 1:**
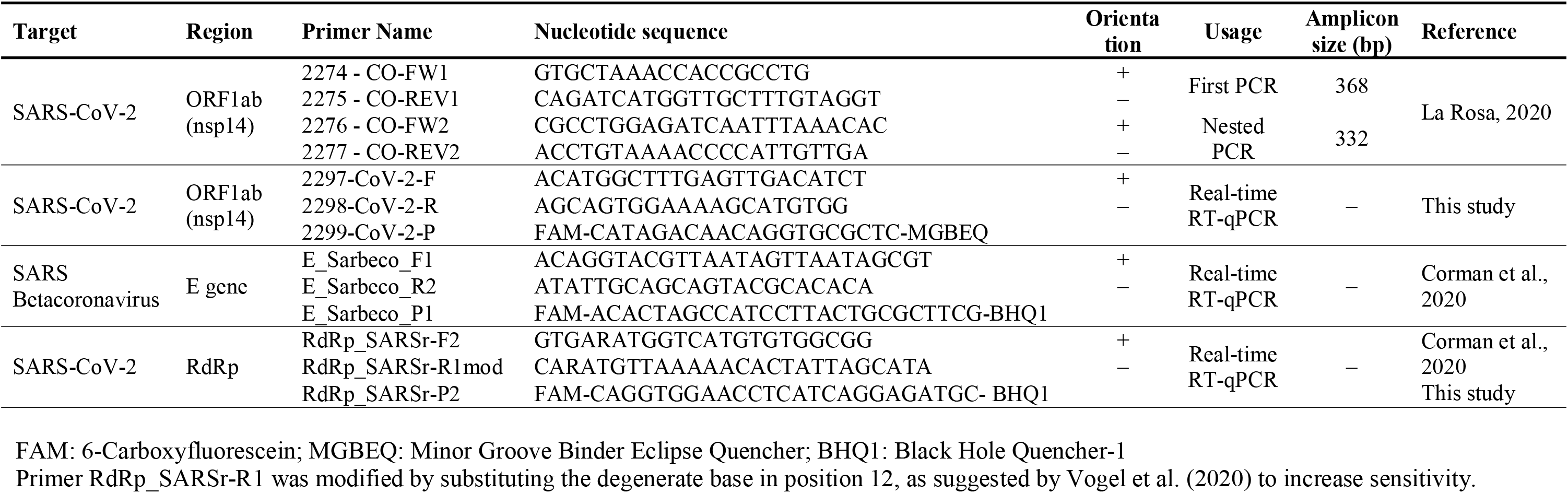
Primers and amplification protocols used in the study.

For the assay, first-strand cDNA was synthesized using Super Script IV Reverse Transcriptase (ThermoFisher Scientific) with the reverse primer, according to the manufacturer’s instructions. PCR reaction was performed using 2.5 μl of cDNA in a final volume of 25 μl (Kit Platinum SuperFi Green PCR Master Mix, Thermo), using 1 μl of the primer (10 μM). The PCR conditions were as follows: 98 °C for 30 sec; 35 cycles at 98 °C for 10 sec, 54 °C for 10 sec, and 72 °C for 30 sec; final extension at 72 °C for 5 min. After the first round of PCR, nested PCR was performed using 2 μl of the first PCR product under the same conditions. A synthetic DNA fragment (Biofab Research, Italy) including the PCR target region was used as positive control. To avoid false-positive results, standard precautions were taken and results were confirmed in two independent experiments.

The PCR products were visualised by gel electrophoresis, were purified using a Montage PCRm96 Microwell Filter Plate (Millipore, Billerica, MA, USA), and were then sequenced on both strands (BioFab Research, Rome, Italy). Sequences were identified using BLAST analysis (https://blast.ncbi.nlm.nih.gov/Blast.cgi). For comparison purposes, all Italian SARS-CoV-2 genome sequences available at the time of analysis (12^th^ June 2020; n=134) were retrieved from Gisaid (https://www.gisaid.org/) and aligned with the study sequences using the MEGA X software (Kumar *et al*., 2018). Sequences were submitted to NCBI GenBank with the following accession numbers: [*a.n. to be assigned*].

### Real-time RT-(q)PCR

Analysis by real-time RT-(q)PCR was undertaken with three different protocols (Table 1):

a. Two published real-time RT-qPCR assays targeting the E gene of the SARS Betacoronavirus and the RdRp gene of SARS-CoV-2, respectively, as described previously (Corman *et al*., 2020) with slight modifications. The RT-qPCR mix (25 μl total volume) was prepared using the UltraSense one-step qRT-PCR System (Life Technologies, CA, USA), and 5 μl aliquots of sample RNA were analysed in reactions containing 1× buffer, 0.1× ROX reference dye, and 1.25 μl of RNA UltraSense enzyme mix. Primer/probe concentrations were as follows: 400 nM, 400 nM and 200 nM for E_Sarberco_F1, E_Sarberco_R2, and probe E_Sarberco_P1, respectively, and 600 nM, 800 nM, and 250 nM for RdRp-SARSr-F2, RdRp-SARSr-R1mod, and probe RdRp-SARSr-P2, respectively. Amplification conditions included reverse transcription for 30 min at 50 °C, inactivation for 5 min at 95 °C and 45 cycles of 15 s at 95 °C and 1 min at 58 °C. For standard curve construction, the two targeted regions were synthetized and quantified by Eurofins Genomics (Germany). Tenfold dilutions were used for standard curve construction.
b. A newly developed real-time RT-(q)PCR designed using the Primer3 software (http://primer3.ut.ee/) targeting the ORF1ab region (nsp14; 3’-to-5’ exonuclease) of the SARS-CoV-2 genome (positions 18600-18699 of GenBank accession number NC_045512). Following optimization, the RT-qPCR mix (25 μl total volume) was prepared using the AgPath-ID One-Step RT-PCR (Life Technologies), and 5 μl of sample RNA were analysed in reactions containing 1× RT-PCR buffer, 1 μl of RT-PCR enzyme mix, 1.67 μl of detection enhancer, and 500 nM, 900 nM, and 250 nM of primer 2297-CoV-2-F, primer 2298-CoV-2-R, and probe 2299-CoV-2-P, respectively. Amplification conditions were: reverse transcription for 30 min at 50 °C, inactivation for 5 min at 95 °C and 45 cycles of 15 s at 95 °C and 30 s at 60 °C. For standard curve construction, the targeted region was synthetized and purified by BioFab Research (Italy), and was quantified by fluorometric measure (Qubit, Thermo Scientific). Tenfold dilutions were used for standard curve construction. In vitro synthetized RNA containing the target region was used as an external amplification control to check for PCR inhibition.

Reactions for quantitative analysis were performed in duplicate. Amplifications were considered acceptable if inhibition was ≤ 50% and if standard curves displayed a slope between −3.1 and −3.6 and a R^2^≥ 0.98. All amplifications were conducted on a Quant Studio 12K Flex instrument (Thermo Scientific). Molecular biology grade water served as the no-template control; two negative controls were included in each run to check for reagent contamination and for environmental contamination, respectively.

Since analysis on environmental matrices may occasionally display high fluorescence background or non-exponential amplification (fluorescence ‘drift’) during amplification, a conservative approach was applied for data analysis. All amplification plots were visually checked for exponential amplification, the threshold was manually set at the midpoint of the exponential phase, and a *Cq* cut-off value of 40 was applied to all results.

### Specificity and sensitivity of nested RT-PCR and real-time RT-(q)PCR

Our in-house nested RT-PCR was evaluated for specificity using the European Virus Archive – EVA GLOBAL (EVAg) panel, kindly provided by the Erasmus University Medical Center (Rotterdam, The Netherlands), and consisting of RNAs from different Alfa- and Beta-coronaviruses (HCoV-NL63, HCoV-229E, HCoV-OC43, MERS-CoV, SARS-CoV and SARS-CoV-2). Moreover, all amplicons obtained by nested PCR were sequenced for confirmation and compared with those available in GeneBank and in Gisaid (https://www.gisaid.org/). The real-time RT-(q)PCR was evaluated for specificity using the GLOBA (EVAg) panel and, in addition, in order to exclude possible aspecific signals, specificity was also tested against a panel of nucleic acids from viruses (*n*=32) and bacteria (*n*=15), as detailed in Supplementary Material. Further to this, to assess specificity of the test on samples representative of the natural microbiota of sewage, 24 ‘blank’ sewage samples (see above) were tested by both molecular methods.

As for sensitivity, in the absence of certified reference material for quantitative assays, SARS-CoV-2 RNA provided in the EVAg panel (quantified ∼3×10^4^ genome copies (g.c.)/μl using our in-house real-time RT-(q)PCR) was used to prepare a serial dilution to assess the sensitivity of the method’s on pure samples of target RNA. The same dilutions were then used to spike nucleic acids extracted from SARS-CoV-2 negative sewage concentrates, in order to evaluate the method’s performance in wastewater samples. The dilutions were tested by nested RT-PCR (one replicate) to determine the lower detectable concentration of the method, and were analysed in quadruplicate to calculate the limit of detection (LOD_50_) and the limit of quantification (LOQ) of the real-time RT-(q)PCR assay. LOD_50_ was calculated according to Wilrich and Wilrich (2009), using the tools available in https://www.wiwiss.fu-berlin.de/fachbereich/vwl/iso/ehemalige/wilrich/index.html). LOQ was calculated as the last dilution level at which the relative repeatability standard deviation (RSDr) of the measurements was below 25 % (Hougs *et al*., 2017).

## RESULTS

Our nested RT-PCR was able to detect the presence of SARS-CoV-2 RNA in spiked sewage samples in a concentration of 3.71 g.c./μl. On pure samples of target RNA, the real-time RT-(q)PCR yielded a LOD_50_ of 0.41 g.c./μl (2.05 g.c./reaction) and a LOQ of 3.71 g.c./μl; in sewage samples, LOD_50_ and LOQ were 1.46 g.c./μl RNA (7.30 g.c./reaction) and 7.35 g.c./μl, respectively. As regards the specificity of the two assays, amplification was obtained only in reactions containing SARS-CoV-2 RNA (EVAg Coronavirus panel), and no aspecific amplification was detected for the other human coronaviruses, for the RNA/DNA panel of enteric viruses and bacteria, or for the 24 ‘blank’ sewage samples collected between September 2018 and June 2019.

The recovery efficiency of the concentration and extraction procedure, evaluated with seeded experiments performed in quadruplicate, using the Alphacoronavirus HCoV-229E (ATCC VR-740) showed an average recovery of 2.04 ± 0.70%. Sample inhibition, assessed by real-time RT-(q)PCR, ranged from null to 49.0%, with a median value of 3.2%.

With regard to the 40 sewage samples collected between October 2019 and February 2020 from the WTPs in Milan, Turin and Bologna, SARS-CoV-2 RNA was detected by nested RT-PCR in 18 samples (amplicon sequences confirmed as SARS-CoV-2 by blast analysis) and in 26 samples by the newly developed real-time RT-(q)PCR (Table 2), with an overall agreement between the two assays of 65.0% (26/40 paired results). In 15 samples, SARS-CoV-2 RNA was detected by both methods. Only these samples, that tested positive by both nested and real-time PCR, were considered as confirmed positive samples. None of the samples tested positive using the previously published SARS-CoV-2 RdRp and Sarbeco E gene protocols.

**Table 2:**
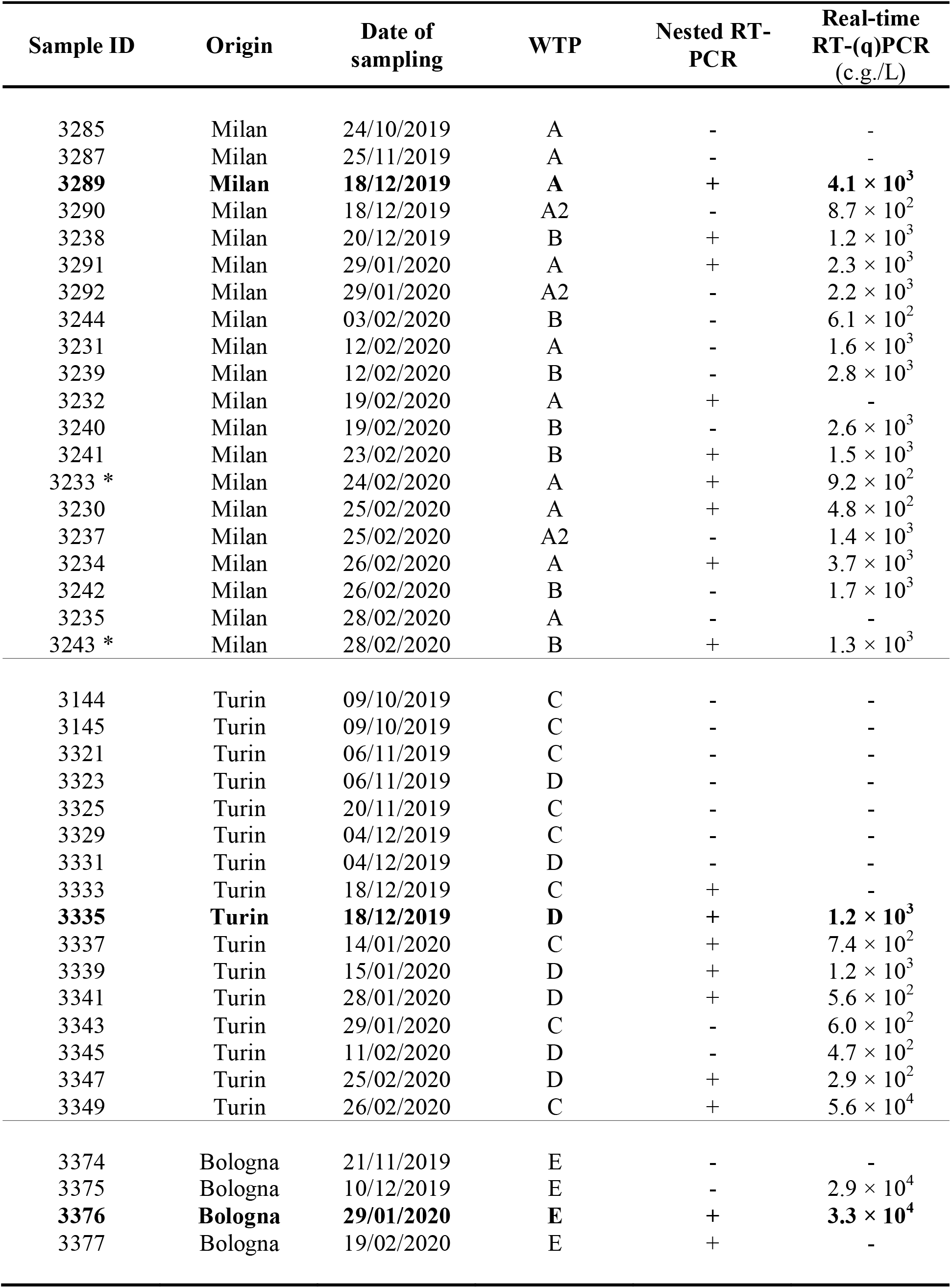
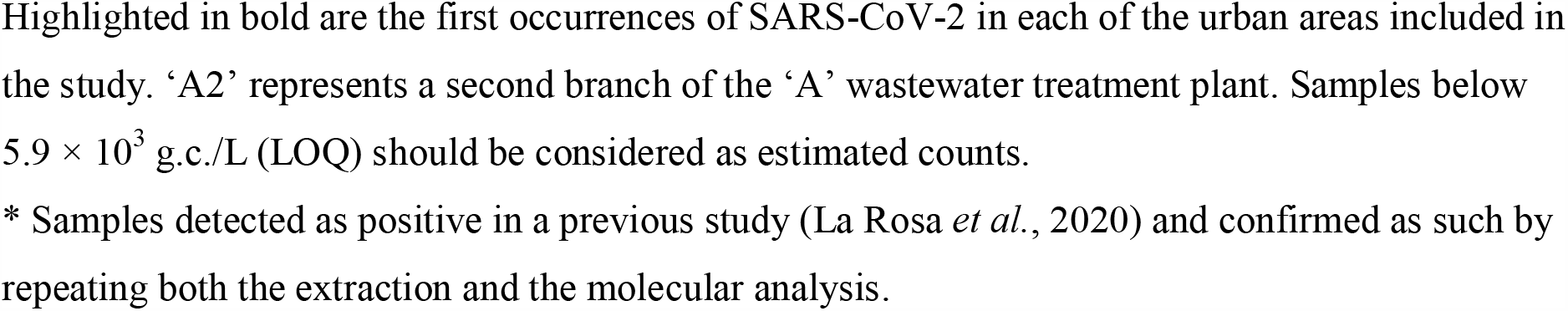
SARS-CoV-2 detection in sewage samples, October 2019 – February 2020.

| Sample ID | Origin | Date of sampling | WTP | Nested RT-PCR | Real-time RT-(q)PCR (c.g./L) |
| --- | --- | --- | --- | --- | --- |
| 3285 | Milan | 24/10/2019 | A | - | - |
| 3287 | Milan | 25/11/2019 | A | - | - |
| **3289** | **Milan** | **18/12/2019** | **A** | + | **$4.1 \times 10^3$** |
| 3290 | Milan | 18/12/2019 | A2 | - | $8.7 \times 10^2$ |
| 3238 | Milan | 20/12/2019 | B | + | $1.2 \times 10^3$ |
| 3291 | Milan | 29/01/2020 | A | + | $2.3 \times 10^3$ |
| 3292 | Milan | 29/01/2020 | A2 | - | $2.2 \times 10^3$ |
| 3244 | Milan | 03/02/2020 | B | - | $6.1 \times 10^2$ |
| 3231 | Milan | 12/02/2020 | A | - | $1.6 \times 10^3$ |
| 3239 | Milan | 12/02/2020 | B | - | $2.8 \times 10^3$ |
| 3232 | Milan | 19/02/2020 | A | + | - |
| 3240 | Milan | 19/02/2020 | B | - | $2.6 \times 10^3$ |
| 3241 | Milan | 23/02/2020 | B | + | $1.5 \times 10^3$ |
| 3233 * | Milan | 24/02/2020 | A | + | $9.2 \times 10^2$ |
| 3230 | Milan | 25/02/2020 | A | + | $4.8 \times 10^2$ |
| 3237 | Milan | 25/02/2020 | A2 | - | $1.4 \times 10^3$ |
| 3234 | Milan | 26/02/2020 | A | + | $3.7 \times 10^3$ |
| 3242 | Milan | 26/02/2020 | B | - | $1.7 \times 10^3$ |
| 3235 | Milan | 28/02/2020 | A | - | - |
| 3243 * | Milan | 28/02/2020 | B | + | $1.3 \times 10^3$ |
| 3144 | Turin | 09/10/2019 | C | - | - |
| 3145 | Turin | 09/10/2019 | C | - | - |
| 3321 | Turin | 06/11/2019 | C | - | - |
| 3323 | Turin | 06/11/2019 | D | - | - |
| 3325 | Turin | 20/11/2019 | C | - | - |
| 3329 | Turin | 04/12/2019 | C | - | - |
| 3331 | Turin | 04/12/2019 | D | - | - |
| 3333 | Turin | 18/12/2019 | C | + | - |
| **3335** | **Turin** | **18/12/2019** | **D** | + | **$1.2 \times 10^3$** |
| 3337 | Turin | 14/01/2020 | C | + | $7.4 \times 10^2$ |
| 3339 | Turin | 15/01/2020 | D | + | $1.2 \times 10^3$ |
| 3341 | Turin | 28/01/2020 | D | + | $5.6 \times 10^2$ |
| 3343 | Turin | 29/01/2020 | C | - | $6.0 \times 10^2$ |
| 3345 | Turin | 11/02/2020 | D | - | $4.7 \times 10^2$ |
| 3347 | Turin | 25/02/2020 | D | + | $2.9 \times 10^2$ |
| 3349 | Turin | 26/02/2020 | C | + | $5.6 \times 10^4$ |
| 3374 | Bologna | 21/11/2019 | E | - | - |
| 3375 | Bologna | 10/12/2019 | E | - | $2.9 \times 10^4$ |
| **3376** | **Bologna** | **29/01/2020** | **E** | + | **$3.3 \times 10^4$** |
| 3377 | Bologna | 19/02/2020 | E | + | - |

Highlighted in bold are the first occurrences of SARS-CoV-2 in each of the urban areas included in the study. 'A2' represents a second branch of the 'A' wastewater treatment plant. Samples below $5.9 \times 10^3$ g.c./L (LOQ) should be considered as estimated counts. \* Samples detected as positive in a previous study (La Rosa *et al.*, 2020) and confirmed as such by repeating both the extraction and the molecular analysis.

Of the 15 positive samples, 8 were taken earlier than February 21, i.e. before the first autochthonous Italian case was reported. Specifically, the first SARS-CoV-2 positive sewage samples were collected as early as and 18 December 2019 in Milan and Turin and 29 January 2020 in Bologna. In all three cities, the virus was also detected in the samples collected subsequently, in January and February, with only one exception - the February sample from Bologna. Here, however, the negative real-time RT-(q)PCR result may have been affected by the slightly higher-than-usual inhibition in this amplification (16.3%). Virus concentration in the positive samples (Table 2 and Figure 2) ranged from <LOD to 5.6 × 10^4^ g.c./L, and most of the samples (23/26) were below the analytical LOQ (5.9 × 10^3^ g.c./L). The highest concentration was recorded in a sample collected in Turin, in February 2020 (plant C, 5.6 × 10^4^ g.c./L).

**Figure 2:**
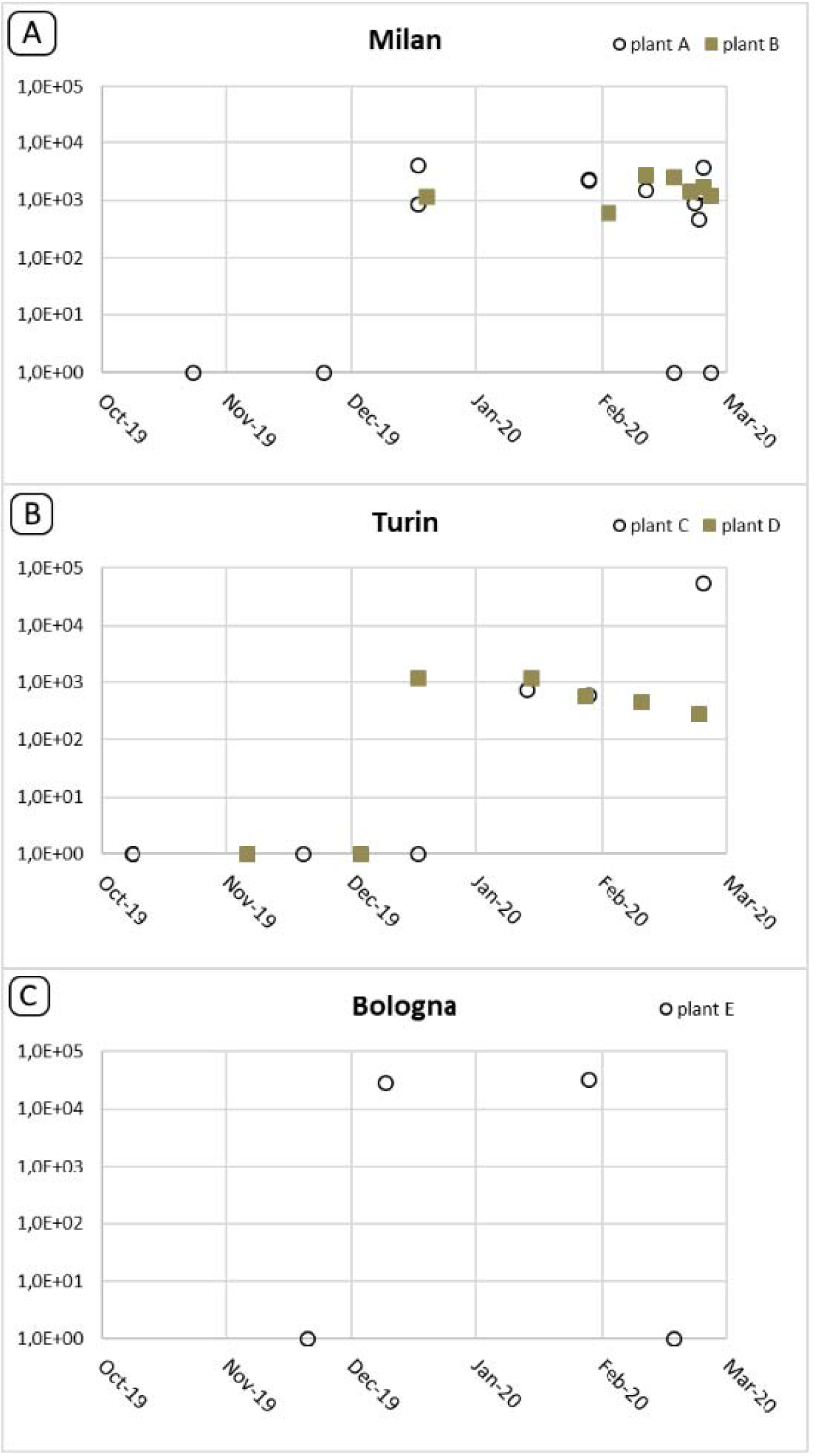
Trend of SARS-CoV-2 detection in Milan, Turin and Bologna during the observed period. All quantitative values obtained by real time RT-(q)PCR are reported, irrespectively of confirmation of positive results by nested RT-PCR

## DISCUSSION

The COVID-19 pandemic first broke out in December 2019 in Wuhan, China, and then rapidly spread worldwide. As of 22 June 2020, 9 million cases of COVID-19 have been registered, and over 470 thousand deaths have been reported (https://www.worldometers.info/coronavirus/).

Italy is one of the first and most severely affected countries in Europe, with a high number of documented cases and deaths. The first documented cases (30^th^ January 2020) were two Chinese tourists who fell ill in Italy in late January after flying in from Wuhan, where the epidemic began. The first autochthonous case of infection was recorded in Italy on 21 February 2020. A sustained local transmission has been documented, so that by 15 June 2020, 237.695 COVID-19 cases had been diagnosed, with 33.168 deaths (https://www.epicentro.iss.it/coronavirus/bollettino/Infografica_15giugno%20ITA.pdf). As far as we know, COVID-19 first affected Lombardy and Veneto and, later on, all the other regions of Italy. The vast majority of cases were reported in Northern Italy. Phylogenetic analyses on SARS-CoV-2 sequences conducted at the beginning of the epidemic, cluster Italian sequences far from the first two Chinese tourists’ strains, and suggest that there may have been multiple introductions of the virus into Italy (Bartolini *et al*., 2020; Giovanetti *et al*., 2020; Stefanelli *et al*., 2020), followed by autochthonous transmission. A genomic characterisation and phylogenetic analysis performed on complete SARS-CoV-2 genomes isolated from patients involved in the first outbreak of COVID-19 in Lombardy, suggested that SARS-CoV-2 entered Northern Italy weeks before the first reported case of infection (Zehender *et al*., 2020).

To test this hypothesis, we analysed sewage samples collected between October 2019 and February 2020 in Northern Italy in the framework of WBE projects on enteric viruses and stored in the archive of the Department of Environment and Health at the Italian National Institute of Health. In a previous study, we demonstrated the presence of SARS-CoV-2 in untreated wastewaters in Italy by analysing samples collected during the early stages of the epidemic (February to April 2020) (La Rosa *et al*., 2020), and other studies around the world have demonstrated that SARS-CoV-2 surveillance in sewage may be considered a sensitive tool to monitor the spread of the virus in the population (Ahmed *et al*., 2020a;Hata *et al*., 2020;Medema *et al*., 2020;Randazzo *et al*., 2020;Wu *et al*., 2020;Wurtzer *et al*., 2020; Kocamemi *et al*., 2020; Bar-Or *et al*., 2020)).

In this study, the analysis of archival samples shows that SARS-CoV-2 was already circulating in Italy, shed by symptomatic, asymptomatic or paucisymptomatic people, many weeks before the first documented autochthonous case, reported on February 21th. Specifically, viral RNA first occurred in sewage samples collected on December 18th, in Milan (Lombardy) and Turin (Piedmont). Therefore, after mid-December 2019, SARS-CoV-2 had already been circulating in major urban centres surrounding the area (Codogno, in the province of Lodi) where the first case of COVID-19 was reported in February 2020. Significantly, all of these regions documented COVID-19 cases starting from 25 February (Protezione Civile, 2020).

A considerable body of evidence supports the hypothesis that SARS-CoV-2 had been circulating in Italy, as well as in other countries, as early as the end of 2019. Indeed, the fact that the virus had been circulating in Europe in late December 2019 has already been demonstrated by a French study (Deslandes *et al*., 2020) that retrospectively analysed samples taken from intensive care patients with influenza-like symptoms in Paris, and found one SARS-CoV-2 positive respiratory sample in a French resident who had not visited China and who had been hospitalised on December 27. Considering the incubation period of COVID-19 - 6.4 days on average (Wang *et al*., 2020) - as well as evidence showing that viral shedding may occur in asymptomatic patients (Jiang *et al*., 2020; Park *et al*., 2020; Tang *et al*., 2020), it is conceivable that the virus was circulating and being released into the sewage in the Paris area roughly at the same time as in northern Italy, as indicated by our positive sewage samples.

Further evidence that SARS-CoV-2 had been spreading far earlier than previously thought came from the United States, where the California health authorities announced that, according to autopsy results, the first death from COVID-19 had to be backdated to 6 February 2020, approximately three weeks before the previously ascertained first US death from the virus (https://edition.cnn.com/2020/04/22/us/california-deaths-earliest-in-us/index.html). In China, where the first case of a novel pneumonia in Wuhan city, Hubei province, was reported in late December 2019, a retrospective analysis identified a patient with symptom onset as early as 1 December (WHO, 2020;Zhang & Holmes, 2020). These indications support scientists’ suspicions that SARS-CoV-2 had been circulating undetected for a relatively long period before the first wave of the epidemic hit.

Additional indications pointing to SARS-CoV-2 circulation before the identification of clinical cases come from WBE studies. In Spain, SARS-CoV-2 was detected in Barcelona wastewaters 41 days (January 15) before the declaration of the first COVID-19 case (February 25), clearly illustrating the ability of wastewater surveillance to anticipate the appearance of cases in the population (Chavarria-Mirò *et al*., 2020). Another Spanish study in the region of Murcia detected SARS-CoV-2 RNA in wastewater before the first COVID-19 cases were declared by the local authorities (Randazzo *et al*., 2020). A similar study conducted in France showed SARS-CoV-2 viral genome in raw sewage before the exponential phase of the epidemic, suggesting that the contamination of wastewaters may occur before any significant appearance of clinical cases (Wurtzer *et al*., 2020).

The hypothesis of SARS-CoV-2 circulation before the identification of the first clinical cases is supported by other epidemiological approaches as well: a seroprevalence study, conducted on healthy blood donors in the province of Milan during the COVID-19 epidemic showed that, at the beginning of the outbreak (24 February), 2.0% of donors displayed IgG for SARS-CoV-2 (Valenti, *et al*., 2020), suggesting that the virus had already been circulating in the population of Milan before the presumed beginning of the outbreak.

Evolutionary sequence analyses lend credibility to the scenario of an introduction of SARS-CoV-2 into the human population in the fourth quarter of 2019 (Duchene *et al*., 2020; Giovanetti *et al*., 2020; Hill & Rambaut, 2020; Li *et al*., 2020;Lu *et al*., 2020; Volz *et al*., 2020). Recently, van Dorp and co-workers analysed the genomic diversity of SARS-CoV-2 in the global population since the beginning of the COVID-19 pandemic by comparing 7666 SARS-CoV-2 genomes covering a vast geographical area (van Dorp *et al*., 2020). Results showed that all sequences shared a common ancestor towards the end of 2019 (6 October 2019 - 11 December 2019), indicating this as the period when SARS-CoV-2 jumped into the human population, and that the virus may have been transmitted between human hosts for quite some time before it was identified. Other elements in support of our findings may be found in the press. According to a Reuters report from March 26, a “significant” increase in the number of people hospitalised with pneumonia and flu-like symptoms in the areas of Milan and Lodi had been documented between October and December 2019 (https://it.reuters.com/article/idUSKBN21D2IG). Clearly, since the virus was still unknown on those dates, the disease would likely have been diagnosed as flu-related. In Turin, between December 2019 and February 2020, the number of patients with a chest CT-scans consistent with COVID-19 pneumonia was four times higher than the number of retrospectively examined CT-scans between December 2018 and February 2019 (https://torino.repubblica.it/cronaca/2020/06/19/news/boom_di_polmoniti_invernali_gia_a_dicembre_il_coronavirus_circolava_a_torino-259645769/). In the region of Liguria, a study conducted by the regional health services (ALISA) showed that samples from blood donors collected in early January revealed the presence of anti-SARS-CoV-2 IgG, thus pointing to an infection in December (https://www.news1.news/en/2020/05/the-coronavirus-arrived-in-liguria-long-before-codognos-patient-zero-2.html).

In agreement with the above data, our study indicates that SARS-CoV-2 was present in Italy before the first imported cases were reported in late January 2020. Since faecal viral shedding occurs in both symptomatic and asymptomatic patients, the question remains whether the traces of SARS-CoV-2 RNA that we found in the sewage of Milan, Turin and Bologna reflected the presence of a significant number of asymptomatic carriers, or of symptomatic patients diagnosed as cases of influenza.

In the present study, several analytical issues had to be addressed. The method used for sample concentration is a modified protocol for the surveillance of poliovirus in sewage. Different volumes and concentration methods are being applied in the various studies assessing the occurrence of SARS-CoV-2: adsorption-extraction with different pre-treatment options, centrifugal concentration device methods, polyethylene glycol concentration, and ultrafiltration (Ahmed et al., 2020b). The concentration method used in this study, based on the two-phase (PEG-dextran) separation method, was selected despite the fact that recovery efficiencies seem to be lower than those obtained by other methods (Ahmed et al., 2020b). It is, however, recommended by the WHO Guidelines for environmental surveillance and is the standard for enteric virus sewage surveillance worldwide (WHO, 2003). This means that a number of laboratories already have both the know-how and the equipment necessary to perform it. Moreover, samples that are routinely collected and concentrated for poliovirus surveillance could be shared and used for SARS-CoV-2 surveillance as well, thus optimising economic and personnel resources.

As for the method used for SARS-CoV-2 detection and quantification, the nested RT-PCR targeting the ORF1ab region, previously published for the first detection of SARS-CoV-2 in wastewater in Italy (La Rosa *et al*., 2020), was tested in this study for specificity against a panel of human coronavirus RNAs and ‘blank’ samples. Moreover, as a routine procedure for all conventional PCRs, the identity of all amplified fragments was confirmed by sequencing. The newly designed real-time RT-(q)PCR assay described in this study was shown to be specific for SARS-CoV-2 by testing against the human coronavirus panel, nucleic acids from relevant viruses and bacteria and ‘blank’ samples. While cross-reactivity with untested microorganisms or with uncharacterised viruses displaying sequences closely matching the target region may not be excluded in principle, the absence of any amplification in ‘blank’ samples seems to confirm the specificity of the reaction. Further tests on a larger variety of reference strains and complex matrices, however, should be performed for full validation of this method. The sensitivity of the real-time RT-(q)PCR assay targeting nsp14 was also assessed, using spiked nucleic acids simulating a wastewater matrix contaminated with SARS-CoV-2. In preliminary tests on these samples this in-house assay proved to be more sensitive than the RdRp test (Corman *et al*., 2020) recommended for the screening clinical samples (data not shown). Indeed, sewage is a very complex matrix, and assays developed for clinical samples are not always suitable for use on environmental samples. It should be noted that, in the absence of an internationally recognised standard for SARS-CoV-2 quantification (as available for other human viruses), a robust assessment of the sensitivity and accuracy of real-time RT-(q)PCR assays cannot be performed, as quantitative results are prone to error depending on both the amplification efficiency of the reactions and the trueness of the reference values attributed to standard curves. Indeed, several studies performing the simultaneous quantification of samples by multiple targets or protocols, as required for example in the CDC protocol testing for N1 and N2 (CDC, 2020), showed significant variability in the values resulting from the different targets (Nemudryi *et al*., 2020, Randazzo *et al*., 2020, Wu *et al*., 2020, Peccia *et al*., 2020, Hata *et al*., 2020), at times displaying differences of up to 3 log in the quantities estimated through different protocols (Chavarria-Miro et al., 2020). Further method harmonization, the development of certified reference materials and a robust characterisation of the method’s performance (including estimation of LOD, LOQ and measurement uncertainty) are required for a reliable use of real-time RT-(q)PCR in SARS-CoV-2 quantification in sewage, particularly in view of the use of these data for WBE, as done in some recent studies (Ahmed *et al*., 2020a).

In this study on samples from the pre-epidemic period (October 2019 to February 2020), virus concentrations in the tested wastewater samples ranged from undetectable to 5.6 × 10^4^ g.c./L, with most results in the order of 10^2^–10^3^ g.c./L. These results are consistent with the concentrations obtained by other authors who tested samples collected at a later stage of the pandemic (mid-January through May 2020) in different countries, finding values ranging from 10^2^ to 10^6^ g.c./L (Ahmed *et al*., 2020a; Chavarria-Mirò *et al*., 2020; Randazzo *et al*., 2020; Wu *et al*., 2020; Wurtzer *et al*., 2020; Kocameni *et al*., 2020). In some of these studies, an upward trend in viral concentrations was observed over the course of the epidemic. Wurtzer et al. (2020) showed SARS-CoV-2 concentrations in Paris wastewaters to increase from 10^4^–10^5^ g.c./L at the beginning of the epidemic to 10^6^–10^7^ g.c./L after its peak. An increase in line with the trend in the local population was also observed in a New Haven (Connecticut, US) study, where concentrations rose from 10^6^ to 10^8^ g.c./L (Peccia *et al*., 2020) and in Barcelona (Spain), where virus amounts went from less than 10^2^ g.c./L at the beginning of the monitoring to approximately 10^4^ g.c./L, and then progressively declined again toward 10^2^ g.c./L (Chavarria-Miró *et al*., 2020). In other studies, perhaps due to shorter periods of observation, an almost constant concentration of SARS-CoV-2 in tested samples was reported following its first detection (Randazzo *et al*., 2020; Hata *et al*., 2020). While the high number of results below the LOQ obtained in our study did not allow for an accurate trend analysis, quantitative data in samples from Milan showed that, following the first occurrence of the virus, an almost constant concentration was reached in sewage samples, while in Turin, the different plants sampled, – serving different districts of the metropolitan area – displayed different tendencies, with a more evident increase in concentrations in plant C. Further studies on samples collected from February 2020 are required to assess the trends in viral concentrations as the epidemic unfolded in the different cities. Moreover, possible differences between WTPs and the areas they serve should be taken into account in future surveillance studies.

In conclusion, our study on archival samples collected before the first autochthonous case was detected confirm that SARS-COV-2 was already circulating in Italy after mid-December 2019, as demonstrated in France by retrospective analysis of stored respiratory samples (Deslandes *et al*., 2020). This study also demonstrates the potential of environmental surveillance as an early warning system capable of alerting public health authorities to the presence of an outbreak in a specific population. The activation of national WBE networks for the monitoring of SARS-CoV-2 could contribute to the early detection of a possible second wave of infection, so as to quickly coordinate and implement mitigation interventions, and could establish a surveillance system ready to operate in case of future epidemic events.

## Data Availability

Data Availability

## Acknowledgments

This publication was supported by the European Virus Archive Global (EVA-GLOBAL) project, which provided the Coronavirus RNA panel for the assessment of our in-house PCR. The EVA-GLOBAL project, in turn, received funding from the European Union’s Horizon 2020 research and innovation programme under grant agreement No 871029.

We also wish to acknowledge Prof. Annalaura Carducci of the University of Pisa for kindly providing the HuCoV 229E (ATCC VR-740) strain and Dr. Loredana Cozzi (Istituto Superiore di Sanità, Department of Food Safety, Nutrition and Veterinary public health) for virus replication. We thank the personnel of the integrated water service for providing wastewater samples.

## SUPPLEMENTARY MATERIAL

**Table 1:**
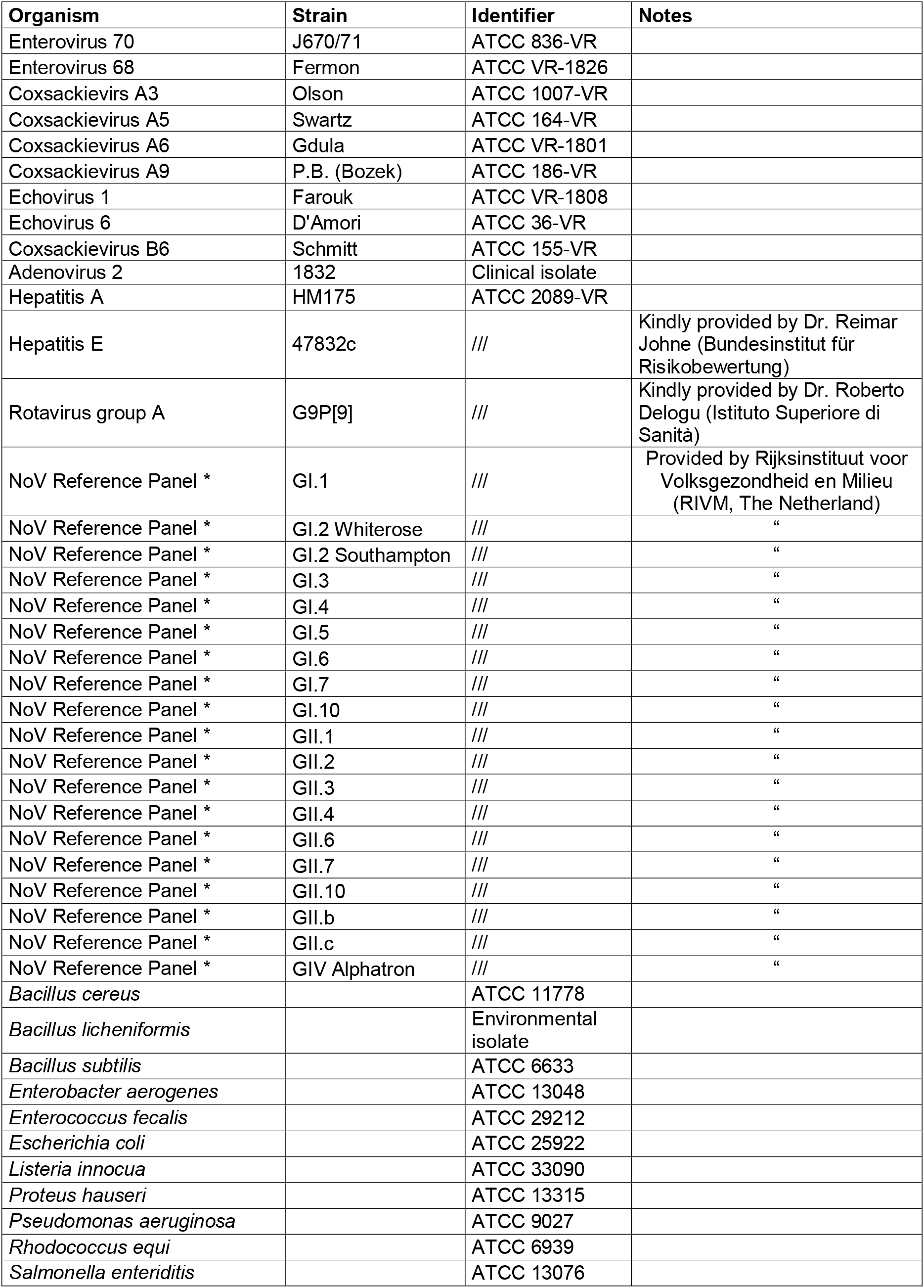
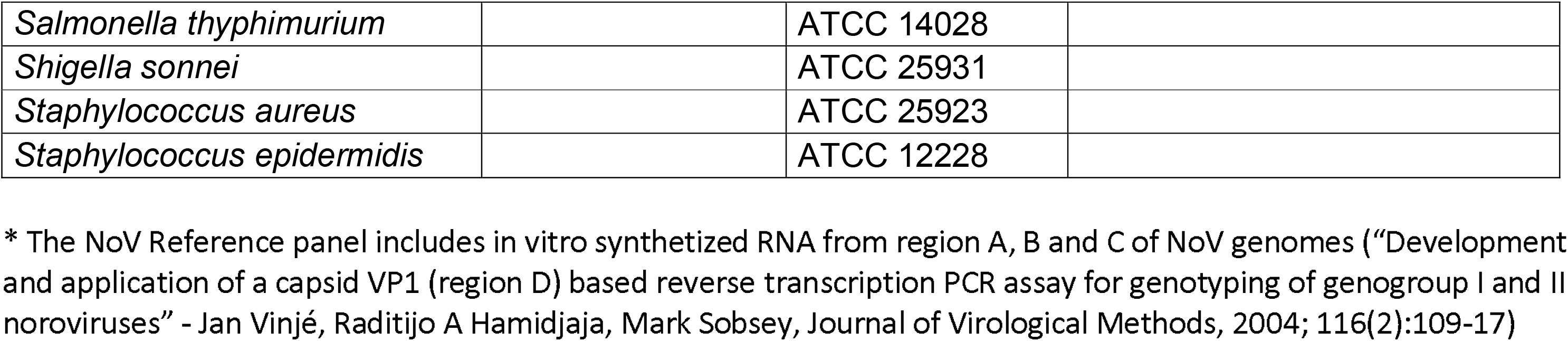
Microorganisms/Nucleic acids tested for evaluation of real-time RT-(q)PCR specificity.

